# ColoSTAT^®^: a multiplexed protein blood test to support colonoscopy triage in patients with symptoms of colorectal cancer

**DOI:** 10.64898/2026.11.19.26368303

**Authors:** Sandhya Nair, Daniel Payne, Dulith De Silva, Orr Zaacks, Erika Spaeth, David Atkins, Andy Feber

**Affiliations:** Rhythm Biosciences Limited, Melbourne, Australia

**Keywords:** biomarkers, early detection of cancer, colonoscopy, colorectal cancer, triage

## Abstract

**Introduction:** Most colorectal cancers (CRCs) are detected after symptomatic presentation in primary care. Because CRC shares symptoms with benign disorders, managing the risk of a missed diagnosis is challenging, and many patients are referred unnecessarily for urgent colonoscopy. As a result, colonoscopy resources cannot keep pace with increasing demand. Simple triage tools that can help prioritise patients for referral are urgently needed.

**Methods:** ColoSTAT^®^ measures the concentration of five protein biomarkers in a single multiplexed ELISA and was validated in a case-control study enriched for cancer cases. Serum samples from two prospective cross-sectional studies and retrospective biobanked samples were combined into training (300) and validation (200) cohorts, matched for sex, age and clinical diagnosis. Overall, 56% of participants were male, and the median age at sampling was 64 years (range, 28–91 years). The validation cohort was supplemented with an additional 100 serum samples from patients with advanced adenoma (AA) (50.0% female; median age 63 years, range 44–84 years).

**Results:** Compared with colonoscopy, ColoSTAT^®^ detected CRC of all stages with a sensitivity of 89% and a specificity of 51% (90.3% and 54.3% in patients aged 45–84 years). Modelled at an intended-use CRC population prevalence of 4%, the negative predictive value (NPV) was >99% and the positive predictive value (PPV) was approximately 7%. Sensitivity was 90.7% for early-stage and 89.4% for late-stage cancers, and 58% for AA.

**Discussion:** ColoSTAT^®^ is a potential blood-based aid for the triage of patients with symptoms of CRC. By helping to prioritise symptomatic patients according to their CRC risk, it could improve triage to colonoscopy and support more effective patient management.

## Introduction

Australia and New Zealand have some of the highest rates of colorectal cancer (CRC) in the world^1^. In Australia, CRC is the fourth most commonly diagnosed cancer and the second leading cause of cancer-related death^2,3^. Colonoscopy remains the gold standard for diagnosing CRC; more than 900,000 procedures are performed annually in Australia, and around 15,500 new CRC cases are diagnosed each year^4^. With up to 50% of colonoscopies returning a normal finding, this low detection rate, combined with the high cost of the procedure (an average of $3200 AUD per colonoscopy^5^), means that CRC poses the highest economic burden of any cancer on the Australian healthcare system, with an estimated annual cost exceeding $1 billion AUD ^6–9^.

Despite active screening programs, most CRCs worldwide are diagnosed after symptomatic presentation to primary care^10^. Only 10–15% of bowel cancers are diagnosed through national screening programs such as Australia’s National Bowel Cancer Screening Program (NBCSP)^4^. In Australia, for example, of the 15,500 CRCs diagnosed each year, the majority (13,000) are diagnosed following symptomatic referral from primary care to colonoscopy^2,3^. Similar figures are observed in England where, although screening participation is significantly higher, screen-detected cancers still account for only 12% of colorectal cancers diagnosed^11,12^.

Colonoscopy services worldwide are under strain. With the rise in early-onset bowel cancer, increased awareness and an ageing population, colonoscopy referrals are expected to increase by 7–10% annually to 2030, doubling from current levels by 2040^13^. Practitioners, healthcare systems and patient advocacy groups are concerned that rising numbers of colonoscopy referrals will lead to longer waiting times, later diagnosis, worse outcomes and prolonged patient distress. With few formal strategies in place to improve triage to colonoscopy^13,14^, there is a clear need for simple, minimally invasive and affordable tests that can better triage patients and reduce the burden of colonoscopy.

Colonoscopy is considered the gold standard because it can examine the entire colon and remove precancerous polyps during the same procedure. However, it is invasive, carries risks such as bowel perforation and bleeding, requires bowel preparation, and is resource-intensive and expensive. There is therefore considerable interest in alternative tests that can identify patients and triage them to colonoscopy more efficiently.

Bowel cancer screening programs worldwide have adopted faecal occult blood tests (FOBT), including the faecal immunochemical test (FIT), as the mainstay of screening. FIT has also been included in triage guidelines for symptomatic patients in some countries to help improve triage to colonoscopy^15^. In the UK, FIT has been embedded directly in the symptomatic referral pathway with the aim of reducing the number of patients referred for colonoscopy^16–18^. Patients presenting with symptoms who do not meet the criteria for immediate urgent referral are offered a FIT to support referral onto the suspected cancer pathway. Despite this, there are still over 700,000 referrals for colonoscopy a year^19^.

There is increasing evidence that people are more willing to participate when offered a blood test^20–22^. This has led to the development of blood-based circulating tumour DNA (ctDNA) tests and of tests combining faecal molecular profiling with FIT. Although these tests have shown advantages over FIT, particularly in the screening setting, they either still require faecal sampling or are expensive, and are unlikely to be cost-effective in most healthcare systems^23,24^.

ColoSTAT^®^ measures the abundance of a panel of serum proteins to produce a colorectal cancer ‘fingerprint’. We previously developed a simple single-plex version of the test^25^ and, building on these data, have developed a second-generation multiplexed assay. ColoSTAT^®^ combines protein abundance with a proprietary software-based algorithm to classify patients as having either a high or low likelihood of CRC. As a referral decision-support tool, ColoSTAT^®^ could help GPs to identify and prioritise patients with suspected cancer for further investigation.

ColoSTAT^®^ has potential advantages over faecal and other blood-based tests: it is a rapid, reproducible, low-cost test that can be performed as part of the standard diagnostic assessment, providing an additional option to aid the triage of patients with symptoms of CRC^26^. Here, we report the development and validation of ColoSTAT^®^, a blood-based test for colorectal cancer, in an at-risk population.

## Methods

### Patient samples

The patient cohorts used to develop and validate ColoSTAT^®^ have been described previously, under ethics HREC/18/CALHN/534 ^27^. Tables 1, 2 and 3 describe the characteristics of the training, validation and advanced adenoma cohorts, respectively. To be classified as negative, a patient had to have no findings on colonoscopy or a negative (non-neoplastic) result on histological review. Inclusion criteria and definition of the advanced adenomas are described in Supplementary Information 1 and 2.

**Table 1.** ColoSTAT^®^ model training cohort patient characteristics.

|  | **Cancer** | **Non-Cancer** |
|---|---|---|
| **Population** | 150 | 150 |
| **Stage** |  |  |
| 1 | 29 | - |
| 2 | 39 | - |
| 3 | 52 | - |
| 4 | 22 | - |
| Nx | 8 | - |
| **Age** |  |  |
| <45 | 13 | 13 |
| 45-54 | 24 | 27 |
| 55-64 | 46 | 46 |
| 65-74 | 50 | 50 |
| 75-85 | 15 | 14 |
| 85+ | 2 | 0 |
| **Sex** |  |  |
| M | 89 | 82 |
| F | 61 | 68 |

**Table 2.** ColoSTAT^®^ validation cohort patient characteristics. (n) represents the number of patients aged 45–84 years.

|  | **Cancer** | **Non-Cancer** |
|---|---|---|
| **Population** | 100 | 100 |
| **Stage** |  |  |
| 1 | 22 (20) | - |
| 2 | 24 (23) | - |
| 3 | 35 (32) | - |
| 4 | 16 (15) | - |
| Unknown | 3 |  |
| **Age** |  |  |
| <45 | 6 | 6 |
| 45-54 | 15 | 19 |
| 55-64 | 33 | 27 |
| 65-74 | 32 | 37 |
| 75-85 | 13 | 11 |
| 85+ | 1 | 0 |
| **Sex** |  |  |
| M | 55 | 54 |
| F | 45 | 46 |

**Table 3.** Advanced adenoma (AA) patient characteristics.

|  | **Adenoma** |
|---|---|
| **Population** | 100 |
| **Age** |  |
| <45 | 2 |
| 45-54 | 18 |
| 55-64 | 34 |
| 65-74 | 34 |
| 75-85 | 12 |
| 85+ | 0 |
| **Sex** |  |
| M | 50 |
| F | 50 |

This was a case-control study combining prospective cross-sectional recruitment with retrospective biobanked samples. The design is enriched for cancer cases relative to the intended-use population (symptomatic patients in primary care) and does not represent a consecutive, real-world referral sample.

### ColoSTAT^®^ serum testing

The ColoSTAT^®^ Colorectal Cancer Test Kit (Ǫuansys Biosystems, UT, USA) quantifies the concentration of five protein biomarkers associated with colorectal cancer^25,28^ within a single well of a 96-well plate. Each well also contains a reference control.

Serum samples were analysed according to the manufacturer’s instructions (Ǫuansys Biosystems, UT, USA). Briefly, serum samples stored at −80 °C were thawed before analysis; diluted serum samples (50 µL) were aliquoted into the ColoSTAT^®^ test plate and processed according to the ColoSTAT^®^ Colorectal Cancer Test Kit protocol. Within each assay plate, a set of standards was used to generate calibration curves for each biomarker. A negative control (blank) was included for background subtraction, along with two plate ǪC controls to confirm that each ColoSTAT^®^ test plate had run correctly. All samples were imaged using a Ǫ-View™ Imager LS (Ǫuansys Biosystems, UT, USA) to generate optical intensities for each biomarker, which were subsequently pre-processed in Ǫ-View Software (allowing calibration, smoothing, background subtraction and normalisation) to generate a quantitative concentration for each biomarker.

### ColoSTAT^®^ algorithm development

A proprietary diagnostic model was developed using a cohort of 300 patients with known clinical outcomes: histologically confirmed CRC or non-cancer (Table 1). Individual biomarker concentrations from each sample were used to train a logistic regression-based model that generates a cancer “likelihood score”, and to determine the cut-off for classifying samples as cancer positive or cancer negative. The ColoSTAT^®^ model was internally cross-validated using leave-one-out cross-validation (LOOCV) in Python (version 3.11) to generate preliminary values for the area under the receiver operating characteristic curve (AUC), sensitivity and specificity. The final proprietary ColoSTAT^®^ model was subsequently validated in an independent cohort of 300 serum samples (Tables 2 and 3).

During training, the classification cut-off was selected using a pre-specified target sensitivity of 90%. This reflects ColoSTAT^®^’s intended role as a triage tool, in which sensitivity (and the resulting negative predictive value, NPV) determines the safety of a negative result and allows patients to be appropriately deprioritised for colonoscopy. Specificity was not constrained and was whatever value resulted once the sensitivity target was met. The model architecture and cut-off were locked after training and before evaluation in the independent validation cohort, and no adjustments were made during validation.

The sample size was based on the power required to demonstrate the anticipated performance with a pre-specified minimum lower 95% confidence limit. To demonstrate a minimum sensitivity of 85% with 90% power, a minimum of 98 samples was required.

## Results

### Patient characteristics

All patient samples have been described previously by He *et al.*^27^. Briefly, of the 300 serum samples in the training cohort, 150 were from patients with confirmed colorectal cancer (CRC) and 150 from patients without cancer. Men made up 59.3% of the cancer group and 54.7% of the non-cancer group. The median age was 63 years (range 28–91 years) (Table 1).

In the validation cohort, 55% of the 100 patients with confirmed cancer and 54% of the 100 non-cancer patients were male. The median age of the validation cohort was 64 years (range 32–87 years) (Table 2).

The validation cohort was supplemented with an additional 100 patients with advanced adenoma (age range 44–84 years; Table 3, Supplementary Information 1).

Additional clinicopathological features are provided in Table 1 for the training cohort and in Tables 2 and 3 for the validation cohort.

### Development of the ColoSTAT^®^ diagnostic model

In total, 300 age- and sex-matched patients were used to develop the ColoSTAT^®^ model (Table 1). The final proprietary model, using only the five biomarker concentrations, had an AUC of 0.83 (95% CI 0.775–0.875) (Figure 1A).

**Figure 1.**
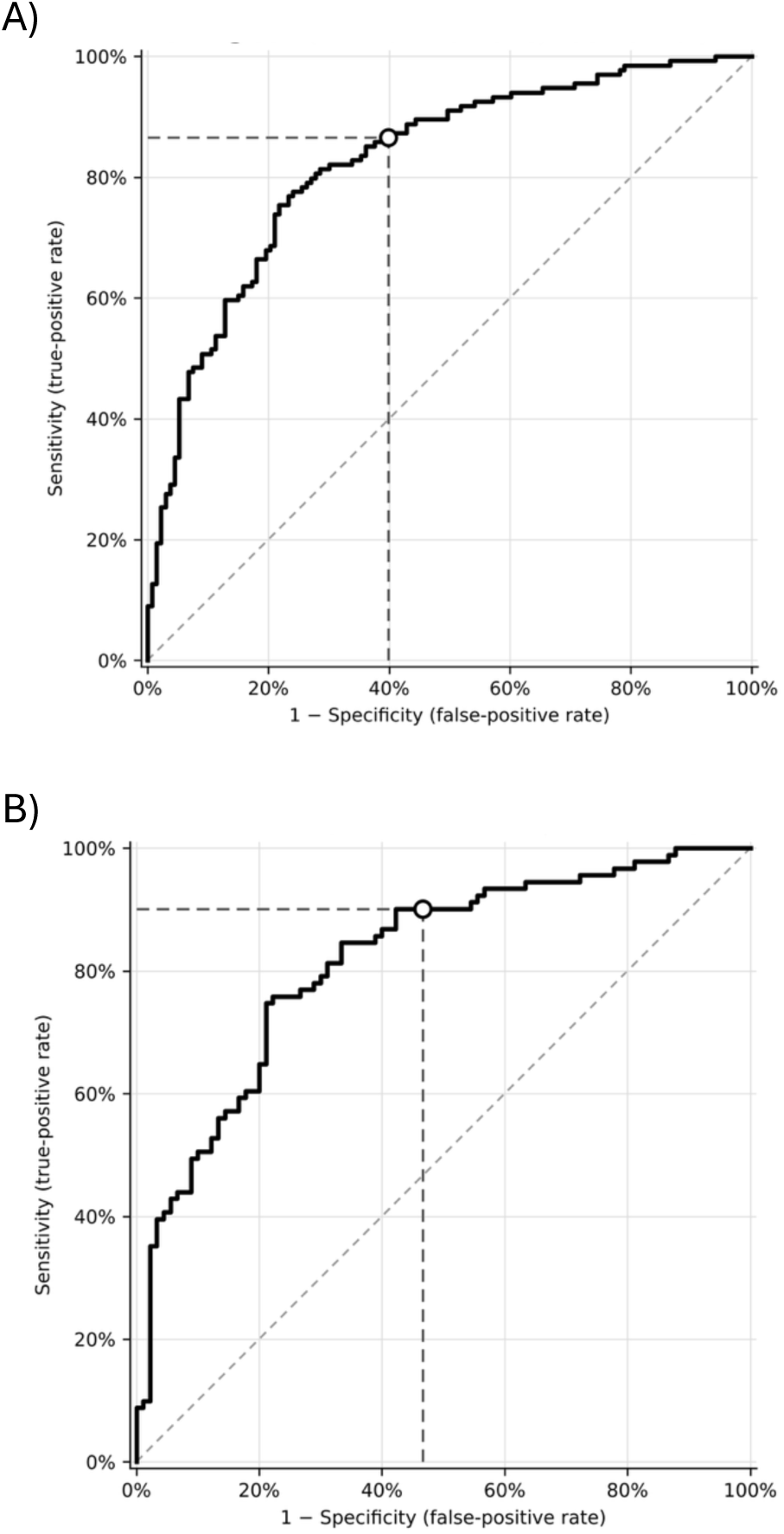
Receiver operating characteristic (ROC) curves for the ColoSTAT^®^ model in (A) the training cohort and (B) the validation cohort. ROC curves for the ColoSTAT^®^ probability score discriminating colorectal cancer (CRC) from non-cancer controls in (A) the training cohort (n = 300) and (B) the independent validation cohort (n = 200). The open circle marks the operating point at the ColoSTAT^®^ classification cut-off, with dashed lines to the corresponding sensitivity and false-positive rate (1 − specificity). The diagonal dashed line indicates no discrimination (AUC = 0.5). AUC, area under the curve.

In the training cohort, ColoSTAT^®^ achieved a cross-validated sensitivity of 85% (95% CI 79.8–91.1) and specificity of 62% (95% CI 54.0–69.4) when trained on 150 known cancers and 150 controls. By stage, ColoSTAT^®^ achieved a sensitivity of 88.2% for stage I/II disease and 81% for stage III/IV disease.

### ColoSTAT^®^ validation performance

ColoSTAT^®^ performance was then validated in 300 independent patient samples, comprising serum from 100 patients with CRC, 100 without cancer and 100 with advanced adenoma (Tables 2 and 3). The AUC for the validation cohort was 0.818 (95% CI 0.757–0.880) (Figure 1B).

Compared with pathological confirmation and including all patients, ColoSTAT^®^ showed an overall sensitivity for CRC of 89% (95% CI 81.4–93.7) and a specificity of 51% (95% CI 41.3–60.6). The prevalence-adjusted NPV of the ColoSTAT^®^ assay was >99% (95% CI 98.41–99.50), modelled using an assumed intended-use population CRC prevalence of 4%; the corresponding positive predictive value (PPV) was 7.6% (95% CI 5.5–9.0%)(Supplementary Table 1).

The ColoSTAT^®^ score was associated with CRC status, with a diagnostic odds ratio (DOR) of 8.4; the odds of CRC increased 2.3-fold (95% CI 1.64–2.52, p < 0.001) per 0.1 increase in ColoSTAT^®^ score.

ColoSTAT^®^ returned a positive result in 58% of patients with advanced adenoma (AA; 58/100). AA was analysed as an outcome separate from the CRC-positive reference standard and was not part of the classifier training labels.

### ColoSTAT^®^ performance by sex and age

Bowel cancer epidemiology differs between males and females, with a higher incidence in males and a higher proportion of right-sided tumours in females^29,30^. We therefore compared both the ColoSTAT^®^ score and detection sensitivity between the sexes. There was no difference between ColoSTAT scores by sex (Mann-Whitney U test: normal p = 0.70, adenoma p = 0.67, CRC p = 0.64; overall p = 0.76). There was also no statistically significant difference in ColoSTAT^®^ detection sensitivity (Fisher’s exact p = 0.75) between males and females (Supp Fig 3).

We subsequently assessed the association between ColoSTAT^®^ score and patient age. The ColoSTAT^®^ score was independent of age (p = 0.49) (Supp Fig 2), and there was no evidence that the relationship with age differed between the non-cancer controls, adenoma and CRC groups (p = 0.53) (Supp Fig 2).

**Figure 2.**
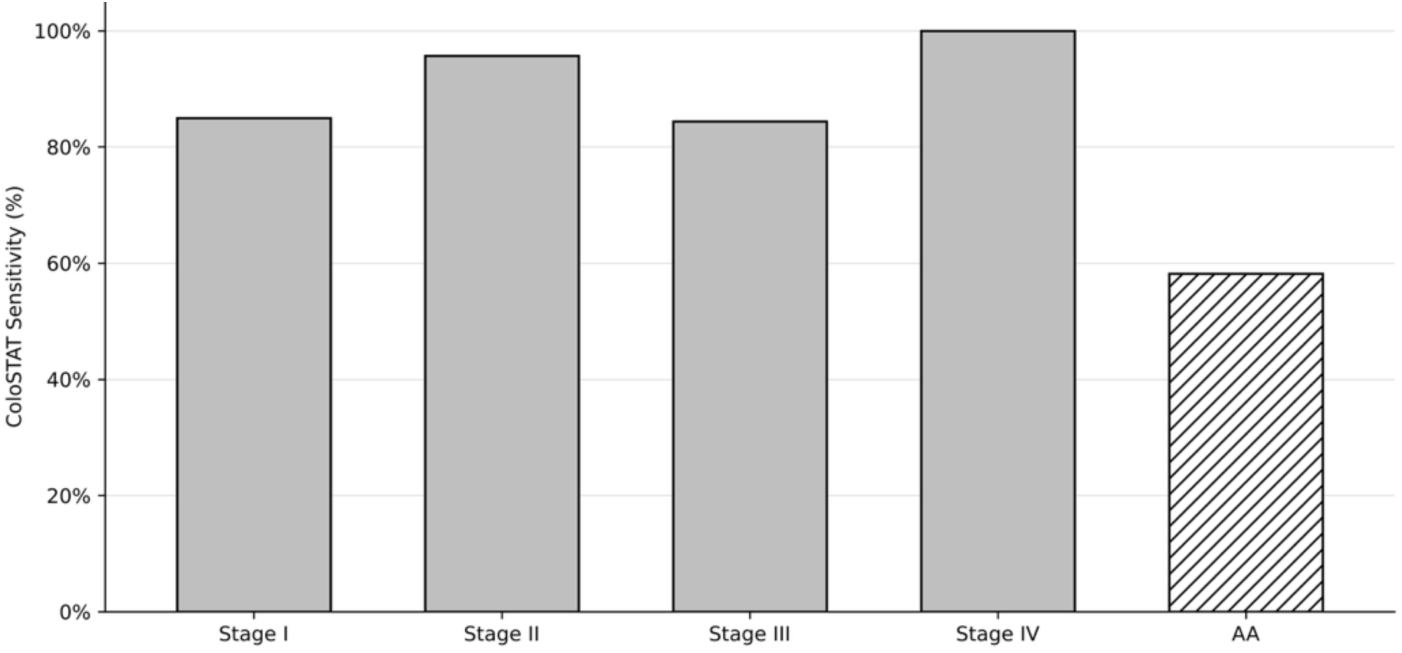
ColoSTAT^®^ sensitivity by CRC stage and positivity in advanced adenoma. Proportion of samples with a positive ColoSTAT^®^ result in patients aged 45–84 years with stage I (n = 20), II (n = 23), III (n = 32) and IV (n = 15) CRC (solid bars), and in patients with advanced adenoma (AA; n = 100; hatched bar).

Patients were stratified into 10-year age intervals, and analysis was restricted to those aged 45–84 years due to the small number of samples from patients younger than 45 (n = 6) or aged 85 and over (n = 1) in the CRC validation cohort. In this age range, sensitivity for CRC was 90.3% and specificity was 54.3%, compared with 89% and 51% in the full validation cohort. Sensitivity for the detection of adenoma was unchanged by the age restriction.

### Early-vs late-stage detection

Early detection is key to improving survival in patients with CRC. A CRC test should therefore detect early-stage CRC with similar sensitivity to late-stage disease, allowing intervention before a cancer progresses. ColoSTAT^®^ was designed to distinguish CRC of any stage from non-CRC; no separate models were built for early- or late-stage CRC, and the proportions of cancers at each stage were similar in the training and validation cohorts (Tables 1 and 2). However, biomarker signals may be stronger in more advanced cancers, so the model could be driven primarily by signal from patients with later-stage CRC, reducing its sensitivity for early-stage CRC.

We explored the ability of ColoSTAT^®^ to detect early (stages I–II) and late (stages III–IV) CRC (Figure 2, Table 4). ColoSTAT^®^ showed similar sensitivity for early-stage (90.7%) and late-stage (89.4%) disease, with no significant difference between the two (Fisher’s exact p = 1.0) (Figure 3, Table 4).

**Table 4.** Sensitivity of ColoSTAT^®^ across CRC stages and advanced adenomas (AA).

| **ColoSTAT Sensitivity (%)<br>Cancer Stage** |  |  |  |  |
| **I** | **II** | **III** | **IV** | **AA** |
|---|---|---|---|---|
| **85.0%** | **95.7%** | **84.4%** | **100.0%** | **58.0%** |

**Figure 3.**
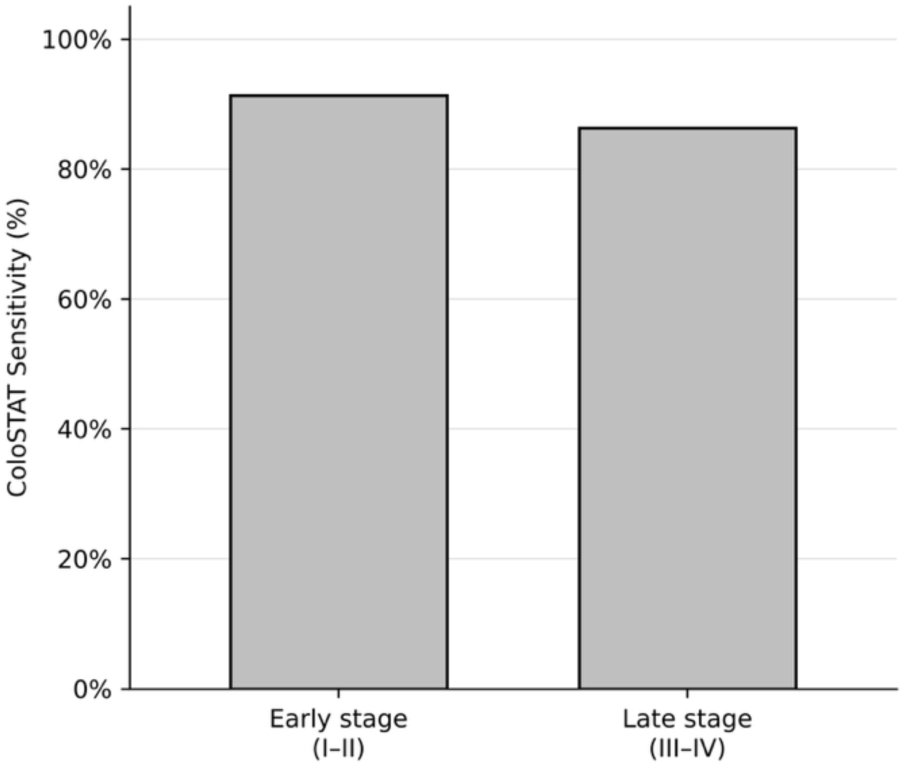
Sensitivity of ColoSTAT^®^ for the detection of early-stage (I–II) and late-stage (III–IV) colorectal cancer in the validation cohort.

The sensitivity of ColoSTAT^®^ did not differ significantly across individual stages (I–IV; Cochran–Armitage p = 0.63, Supp Fig 2B), although there was a non-significant trend towards higher ColoSTAT^®^ scores with increasing tumour stage (p = 0.16; Supp Fig 4).

## Discussion

This study describes the development and validation of ColoSTAT^®^, a five-protein serum test intended to support the triage of patients presenting with symptoms of colorectal cancer. In the validation cohort, ColoSTAT^®^ detected CRC with a sensitivity of 89% and a specificity of 51% (AUC 0.818), and with a sensitivity of 90.3% and a specificity of 54.3% in patients aged 45–84 years. Modelled at a CRC prevalence of 4% (approximating the intended-use population), the negative predictive value (NPV) exceeded 99%. Sensitivity was similar for the detection of early-stage (stage I–II, 90.7%) and late-stage (stage III–IV, 89.4%) disease. Furthermore, ColoSTAT^®^ returned a positive result in 58% of advanced adenomas, although the model was trained on cancer cases only. ColoSTAT^®^ scores were independent of patient age and sex.

These findings suggest that ColoSTAT^®^ may act as a triage-support tool, with its value lying in the high sensitivity and high NPV: a negative result indicates a low residual risk of CRC, which may help clinicians to deprioritise low-risk patients for colonoscopy, while a positive result identifies patients in whom investigation should be prioritised. ColoSTAT^®^ is intended to support, rather than replace, existing referral pathways and clinical judgement.

Referral for colonoscopy based on symptoms alone detects nearly all cancers, but its positive predictive value is low (<7%): most referred patients are not diagnosed with cancer, and many have a normal colonoscopy ^9,31,32^. Rising referral volumes combined with low diagnostic yield are placing colonoscopy services under increasing pressure. Together with capacity constraints, this often lengthens the time to colonoscopy, delays diagnosis and increases the risk of disease progression. Adding a simple triage test to symptom assessment can substantially improve the positive predictive value of symptom-based referral; this approach is already in place in some settings.

In the UK, FIT is embedded in the symptomatic diagnostic pathway (NG12), where a positive FIT result is used as the threshold for referring symptomatic patients for suspected colorectal cancer ^33^. This established pathway illustrates how a complementary blood-based triage test such as ColoSTAT^®^ could be positioned in a routine clinical pathway. ColoSTAT^®^ also offers a blood-based alternative to FIT that does not depend on the collection and return of a stool sample, a recognised barrier to participation^34,20–22^.

Adding ColoSTAT^®^ to the diagnostic pathway, either as an adjunct or an alternative to FIT, could, if confirmed in a prospective clinical-utility study, reduce the number of colonoscopies needed to diagnose CRC effectively and safely. Furthermore, an enhanced, biomarker-led triage strategy for colonoscopy referral could substantially improve patients’ quality of life by reducing the need for an invasive and uncomfortable procedure, the need to take time off work, and the anxiety associated with the procedure and result. Future health economic analyses will be needed to determine the true potential impact of ColoSTAT^®^ on earlier disease detection and the use of downstream resources, together with qualitative analysis of test acceptability to patients and clinicians.

The combination of prospective and retrospective cohorts in the development and validation of the ColoSTAT^®^ model is a potential limitation of this study. Unknown factors, including systematic differences in sample collection, processing, storage or handling between study sites, may have introduced variation between cohorts, particularly as some collections were disproportionately weighted towards cancer samples. The small number of patients younger than 45 years of age also limits our ability to define the utility of ColoSTAT^®^ in younger symptomatic patients, in whom cancer incidence is rising at an alarming rate^35–37^. Patient age was not associated with the ColoSTAT^®^ score (Supp Fig 2), suggesting that ColoSTAT^®^ may perform similarly in patients outside this age range; however, this needs to be confirmed. Future studies will address these limitations through standardised sample collection and the recruitment of patients with early-onset disease.

In conclusion, ColoSTAT^®^ shows potential as a decision-support tool for the triage of symptomatic patients at risk of CRC. In patients with symptoms suggestive of CRC, a low-risk result could help reduce the number of patients referred for urgent colonoscopy, while a high-risk result could identify patients who would otherwise be classed as low risk and prioritise them for colonoscopy, supporting earlier diagnosis. The clinical utility of ColoSTAT^®^ will be validated in large-scale prospective clinical trials.

## Declarations

### Funding

This study was funded by Vision Tech Bio Pty Ltd, a wholly owned subsidiary of Rhythm Biosciences Limited.

### Ethics Approval and Consent

All procedures performed in studies involving human participants were in accordance with the ethical standards of the institutional Human Research Ethics Committees and with the 1964 Declaration of Helsinki and its later amendments. The study was approved by the Central Adelaide Local Health Network Human Research Ethics Committee (reference no. HREC/18/CALHN/534) and endorsed by the Human Research Ethics Committees of the participating study centres.

### Consent to Participate

Informed consent was obtained from all individual participants included in this study. Biobanked samples were obtained from participants who had taken part in an ethics-approved clinical trial conducted according to ICH GCP guidelines and who had provided informed consent, including for their samples to be used in future research.

### Competing Interests Financial

All authors are current or former employees of Rhythm Biosciences Limited, and DA, AF, ES and SN also hold stock in the company.

### Data Availability

**The** study protocol and datasets generated and analysed during this study will be made available on request from.

**Permissions No** material has been reproduced from other sources.

## Author Contributions

SN, DP, AF and DA contributed to the conception, design and planning of the work. SN, DDS and OZ coordinated and performed ColoSTAT^®^ testing. SN, DP and AF performed the overall analysis and interpretation of the study data. AF prepared the manuscript, and DA and ES reviewed and revised it. All authors read and approved the final manuscript.

## Acknowledgments

This research was funded by Vision Tech Bio Pty Ltd, a wholly owned subsidiary of Rhythm Biosciences Limited (Melbourne, Vic, Australia).

---

**Supplementary Figure 1.**
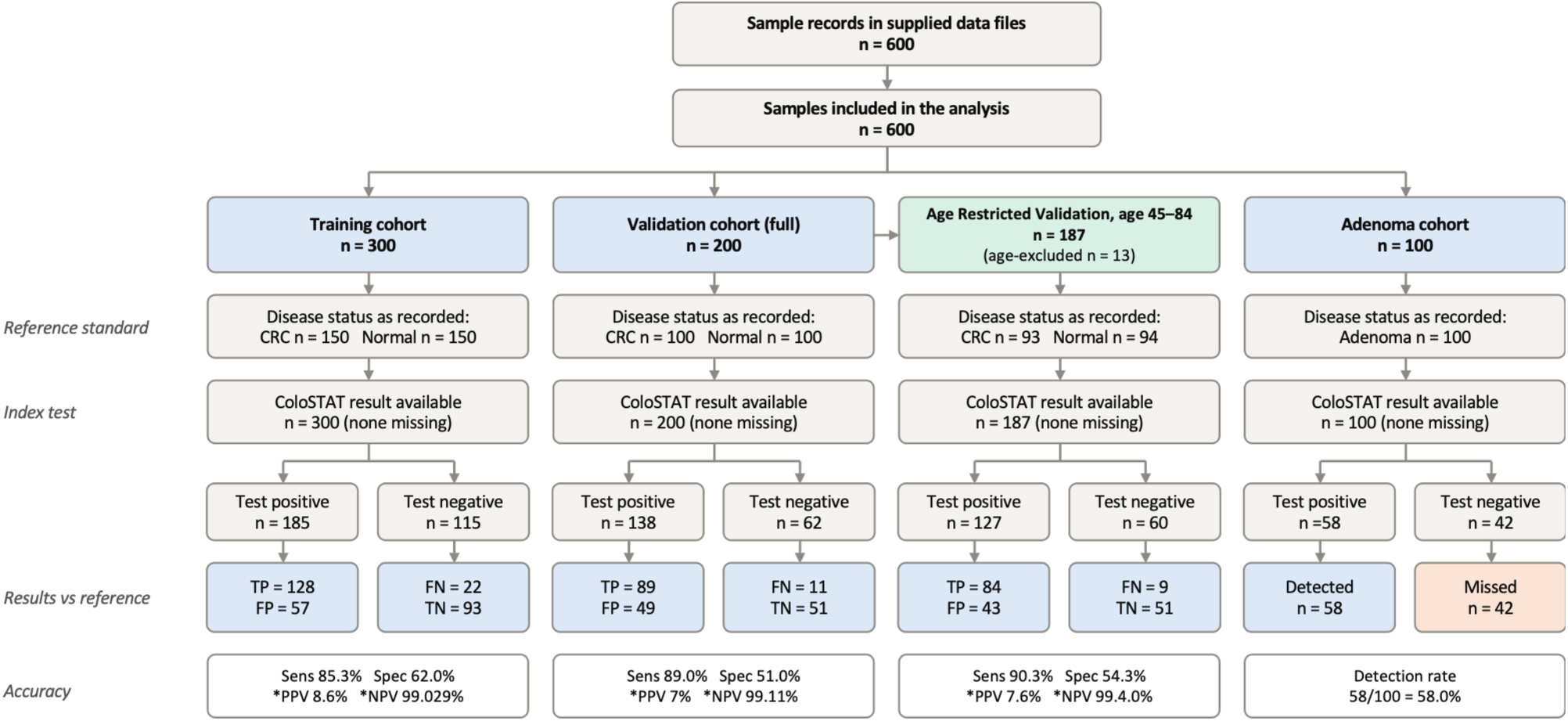
ColoSTAT^®^ STARD flow diagram.

**Supplementary Figure 2.**
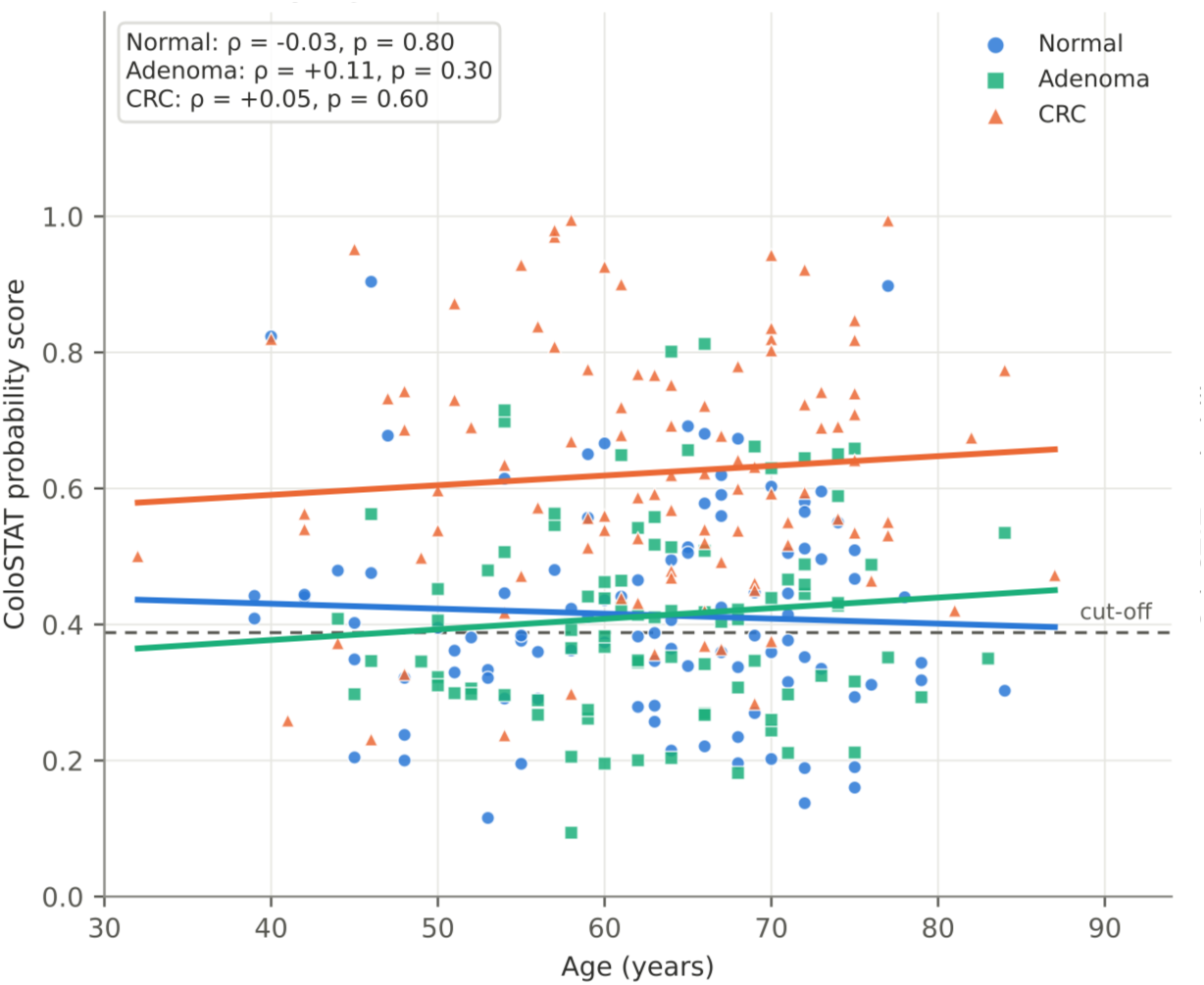
Association of ColoSTAT^®^ score with patient age

**Supplementary Figure 3.**
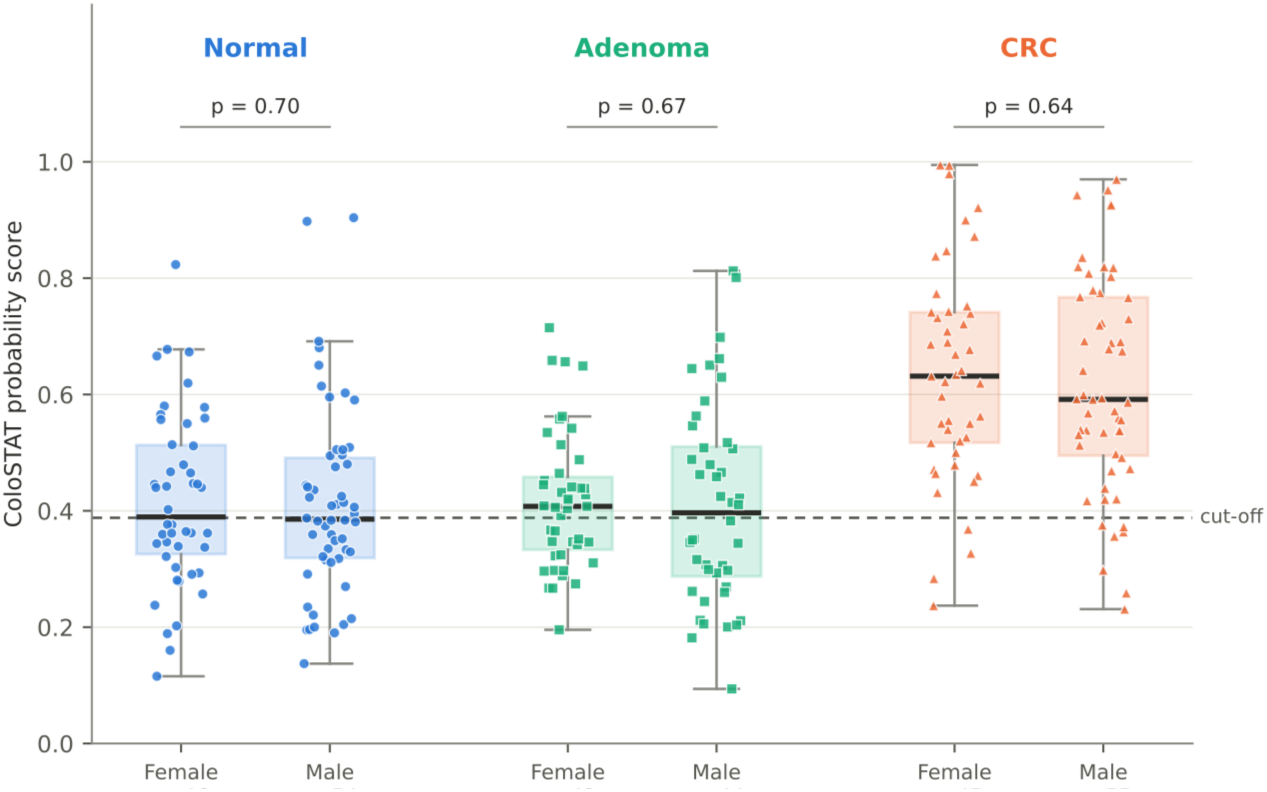
Association of ColoSTAT^®^ score with patient sex

**Supplementary Figure 4.**
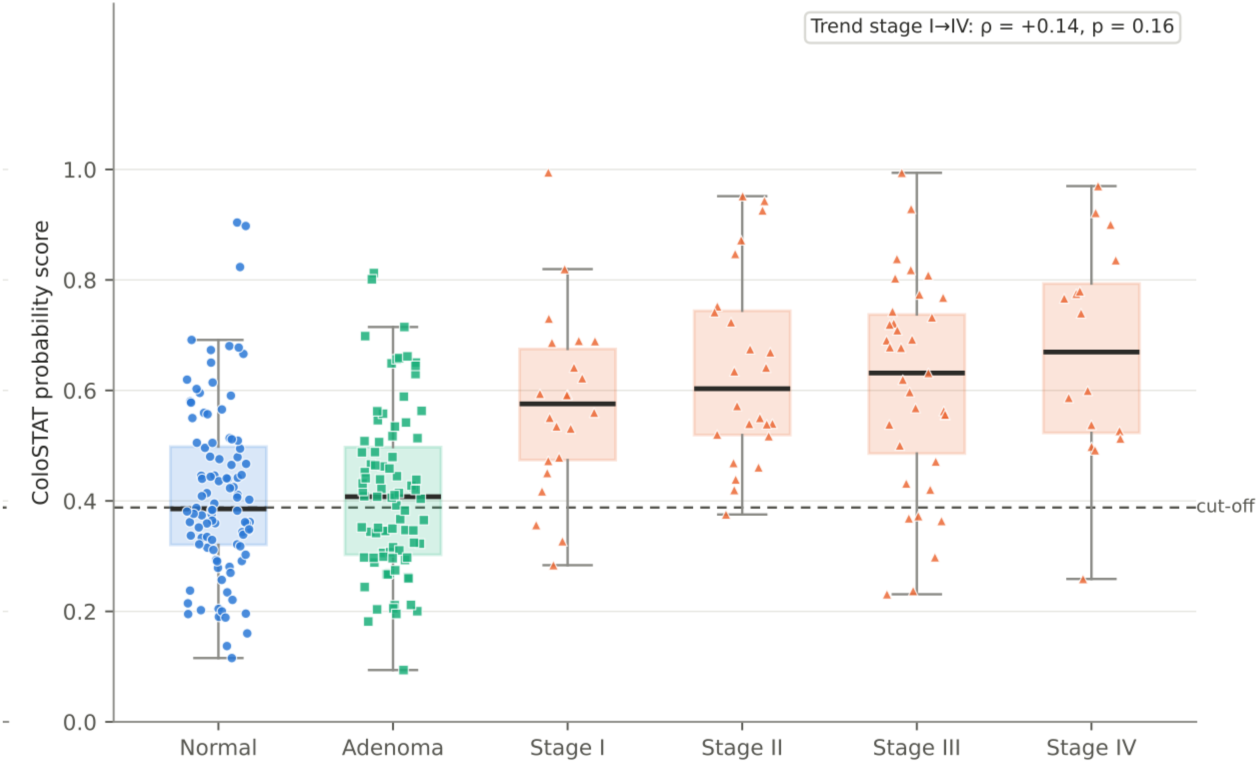
Comparison of ColoSTAT^®^ score by tumour stage.

**Supplementary Table 1.** Observed diagnostic performance of ColoSTAT^®^ in the training (A) and validation (B) cohorts. Two-by-two tables of ColoSTAT^®^ results (positive/negative) against reference-standard colorectal cancer (CRC) status in (A) the training cohort (n = 300; 150 CRC, 150 non-cancer controls) and (B) the independent validation cohort (n = 200; 100 CRC, 100 non-cancer controls). All samples were included, with no exclusions by age. ColoSTAT^®^ results were classified as positive or negative at the pre-specified probability score cut-off. Sensitivity, specificity, positive predictive value (PPV), negative predictive value (NPV), prevalence adjusted PPV and NPV, and accuracy are given as proportions with 95% confidence intervals (CI), calculated by the Wilson score method. PPV and NPV are the values observed in the case-control set, which has a CRC prevalence of 50%.

| **A) Training cohort (n = 300)** |  |  |  |
|---|---|---|---|
|  | **CRC** | **Normal** | **Total** |
| **ColoSTAT positive** | TP = 128 | FP = 57 | 185 |
| **ColoSTAT negative** | FN = 22 | TN = 93 | 115 |
| **Total** | **150** | **150** | **300** |
|  | **n/N** | **% (95% CI)** |  |
| Sensitivity | 128/150 | 85.3 (78.8–90.1) |  |
| Specificity | 93/150 | 62.0 (54.0–69.4) |  |
| PPV | 128/185 | 69.2 (62.2–75.4) |  |
| NPV | 93/115 | 80.9 (72.7–87.0) |  |
| Pa PPV |  | 8.6% (7.0–10.4) |  |
| Pa NPV |  | 99.02% (98.54–99.35) |  |
| Accuracy | 221/300 | 73.7 (68.4–78.3) |  |
| **B) Validation cohort (n = 200)** |  |  |  |
|  | **CRC** | **Normal** | **Total** |
| **ColoSTAT positive** | TP = 89 | FP = 49 | 138 |
| **ColoSTAT negative** | FN = 11 | TN = 51 | 62 |
| **Total** | **100** | **100** | **200** |
|  | **n/N** | **% (95% CI)** |  |
| Sensitivity | 89/100 | 89.0 (81.4–93.7) |  |
| Specificity | 51/100 | 51.0 (41.3–60.6) |  |
| PPV | 89/138 | 64.5 (56.2–72.0) |  |
| NPV | 51/62 | 82.3 (71.0–89.8) |  |
| Pa PPV |  | 7.6% (95% CI 5.5–9.0 |  |
| Pa NPV |  | 99.4% (95% CI 98.41–99.50) |  |
| Accuracy | 140/200 | 70.0 (63.3–75.9) |  |

*TP, true positive; FP, false positive; FN, false negative; TN, true negative; CRC, colorectal cancer; PPV, positive predictive value (observed, non adjusted for prevalence); NPV, negative predictive value (observed, non adjusted for prevalence);Pa PPV value (adjusted for prevalence); Pa NPV value (adjusted for prevalence) CI, confidence interval.*

## Supplementary Information 1

Advanced adenoma was defined as:

Adenoma with carcinoma in situ or high-grade dysplasia, any size;

Adenoma with a villous growth pattern (≥25%), any size;

Adenoma ≥1.0 cm in size; or

Sessile serrated lesions *≥* 1.0 cm in diameter

## Supplementary Information 2

### Patient inclusion exclusion criteria for symptomatic patients

Patients must meet the following criteria:

- Age ≥ 30 years at the time of enrolment.
- Referred for colonoscopy by a GP on clinical grounds with one or more lower gastrointestinal symptoms of CRC, defined as at least one of the following for ≥4 weeks:

- Rectal bleeding (haematochezia or melaena) not attributable to a confirmed haemorrhoidal source
- A change in bowel habit (increased frequency, looser stools, or alternating constipation/diarrhoea)
- Unexplained iron deficiency anaemia (haemoglobin < 120 g/L [females] or < 130 g/L [males] with ferritin < 30 µg/L)
- Abdominal mass on clinical examination
- Unexplained unintentional weight loss ≥5% body weight over 6 months
- Elevated faecal immunochemical test (FIT) result within the past 6 months (quantitative FIT ≥ 10 µg Hb/g)
- Colonoscopy planned to be performed within 90 days of enrolment blood draw.
- Able and willing to provide written informed consent in English or a language for which a validated translation and interpreter service is available.
- Adequate peripheral venous access for a single 10 mL blood draw.

## Notes

### Author Declarations

Central Adelaide Local Health Network Human Research Ethics Committee gave ethical approval for this work.

